# The benefits and risks of maternal RSV vaccination on mortality in South Africa: a modelling study

**DOI:** 10.1101/2025.02.14.25321914

**Authors:** Ayaka Monoi, Akira Endo, Simon R Procter, Sequoia I. Leuba, Stefan Flasche, Mark Jit, Maternal RSV vaccine benefit-risk advisory group, Philippe Beutels, Cheryl Cohen, Daniel R. Feikin, Mihaly Koltai, Shabir A. Madhi, Jocelyn Moyes, Patrick K. Munywoki, Joyce Nyiro, Bryan O. Nyawanda, Erin Sparrow, Heather J Zar

## Abstract

**Background:** Maternal respiratory syncytial virus (RSV) vaccine, RSV prefusion F protein vaccine (RSVpreF (Abrysvo^®^)), was found to be safe and efficacious in its phase III trial. However, post-hoc stratified analyses identified an excess of preterm births in the intervention arm in two upper middle-income countries, most prominently in South Africa. This study weighs the potential benefits and risks in mortality associated with maternal RSV vaccination in South Africa, assuming the increased risk of preterm births observed in the trial was caused by vaccination.

**Methods and Findings:** We compared the estimated RSV-associated infant deaths averted by vaccination (benefits) and neonatal mortality potentially associated with vaccine-associated risk in preterm birth (risks) in South Africa. The benefit model estimated the South African RSV disease burden in 2011-16 and waning vaccine protection during infancy. The risk model estimated excess neonatal mortality using gestational age (GA)-specific mortality data from a South African cohort study and the GA-specific birth distribution in South Africa in the trial, but did not incorporate the mortality risk found in the vaccine trial in which no excess deaths occurred.

The benefit model estimated that vaccination would reduce RSV-associated infant deaths by 31 (95% Credible Interval (Crl): 27, 35) per 100,000 live births born to vaccinated mothers in South Africa. The risk model suggested that neonatal deaths would increase by 44 (95%CrI: −43, 210) with vaccination at 24-36 GA weeks, totaling 1.4 (95%CrI: −1.4, 7.0) excess neonatal deaths for every infant RSV death prevented. Using the data for infants born to mothers vaccinated at 27-36 GA weeks, the predicted risks sharply dropped and in 98% of the simulations the benefits outweighed the risks.

**Conclusions:** If RSVpreF increases preterm birth risk, and if this increases neonatal mortality, then the benefit-risk analysis did not show that the direct benefits of vaccination in reducing RSV-associated infant mortality would substantially outweigh the risks of preterm birth-associated neonatal mortality in South Africa with vaccination from 24 GA weeks. There was large uncertainty in the analyses due to small numbers of preterm births. With vaccination from the third trimester, the benefit-risk analysis favoured vaccination.

## Introduction

Respiratory syncytial virus (RSV) is a significant cause of pediatric morbidity and mortality worldwide, particularly in low- and middle-income countries (LMICs) [3]. It is estimated that globally, RSV leads to approximately 100,000 deaths among children under 5 years in a year [3]. The burden is concentrated in LMICs where over 97% of RSV-attributable deaths occur [3], and especially high among younger infants, who are at the greatest risk of developing severe disease [3–6]. Antillón and colleagues estimated that although children under 6 months of age represent only 10% of the under 5 years population in LMICs, they bear a disproportionate burden of disease, accounting for approximately 30% of hospitalizations and 38%-50% of deaths [5].

Prophylactics against RSV in infancy have recently been licensed, including a maternal RSV vaccine [7] and a long-acting monoclonal antibody [8]. The bivalent RSV prefusion F protein-based vaccine (RSVpreF) developed by Pfizer (Abrysvo^®^, hereafter referred to as “RSVpreF”) has received approval for use in pregnant women from the United States Food and Drug Administration (FDA) and the European Medicines Agency (EMA) following the successful completion of the phase lll MATISSE trial [7] (hereafter referred to as “the trial”). The trial provided evidence of the vaccine’s efficacy in preventing RSV disease in infants, especially severe disease. Mathematical modelling studies suggested that an RSV vaccination programme could reduce RSV-associated mortality and be cost-effective, particularly in LMICs like South Africa [9–11]. However, in the trial, a non-significant imbalance in preterm birth rates was observed: overall this was not statistically significant with 5.7% (95%CI: 4.9, 6.5) of infants being born prematurely in the intervention arm versus 4.7% (95%CI: 4.1, 5.5) in the placebo arm [12]. The imbalance in preterm birth was most pronounced in South Africa [12,13]. However, no imbalance in neonatal deaths between study arms was observed either in the whole trial or in South Africa (e.g., for the whole trial, 8 in the intervention arm vs 14 in the placebo arm with relative risk of 0.57 (95%CI: 0.24, 1.36)) [1,12]. Nevertheless, the observed imbalance in preterm birth rates has led to concerns about vaccine safety. Notably, a trial of another RSVpreF vaccine in pregnant women was terminated due to increased preterm birth rates in the intervention arm, albeit with an associated excess mortality [14]. To inform national decision-making, it is crucial to balance the risks of maternal RSV vaccination against the benefits [15] in the local context.

This study aimed to assess the potential impact of maternal RSV vaccination with RSVpreF in South Africa on mortality, should the excess risk of preterm birth associated with vaccination be substantiated and should preterm births translate into mortality. To this end, we compare the vaccine benefits of reducing RSV-associated infant mortality against the potential risks of increased neonatal mortality due to preterm birth.

## Methods

To assess the risks of RSVpreF associated with the potential safety signal from the South African component of the trial, for the purpose of this analysis we assumed that maternal RSV vaccination is causally associated with an increased risk of preterm birth, which contributes to an increased risk of neonatal mortality. It is important to note that it is currently impossible to assess the true nature of this potential safety signal with any certainty and that we are not trying to do this here, instead we investigate the hypothetical mortality implications if the signal were to be confirmed subsequently.

We quantified the benefits measured as the number of RSV-associated deaths prevented in the first year of life, and the risk as the number of potential excess neonatal deaths associated with preterm birth per 100,000 live births born to South African mothers vaccinated with RSVpreF. The phase III trial birth data used in this study can be downloaded from [1]. The phase III trial efficacy data were obtained from [13]. Data from the Drakenstein Child Health Study [2] were obtained from Heather Zar. Analysis was performed using R version 4.3.3. [16]

### Benefits

Vaccine-preventable RSV-associated infant deaths were estimated using a previously described model of vaccine impact [10] with updated assumptions on vaccine efficacy and its waning. In brief, age-specific South African RSV-associated deaths were estimated in the model, using the baseline RSV incidence from South Africa in 2011-2016. Deaths averted through vaccination during the first year of life were then estimated by multiplying the number of RSV-associated deaths by age-specific vaccine efficacy. We assumed uniform vaccine protection among vaccinees, regardless of their gestational ages [17]. We also assumed that there were no indirect effects of vaccination, i.e. that maternal vaccines do not affect the overall transmission dynamics of RSV beyond the vaccinated mother and her infant.

#### RSV-associated Infant Mortality

RSV-associated infant mortality in South Africa has been estimated in our static cohort model [10]. We first estimated age-stratified RSV-associated severe acute respiratory illness (SARI) rates in South Africa using country-specific surveillance and ecological data in 2011-2016 [6]. We then estimated RSV-associated infant deaths using the SARI hospitalization rates, in-hospital case fatality rate (CFR), and accounted for underreporting due to out-of-hospital deaths estimated from national vital statistics data [18].

To assess the uncertainty in our estimates, we performed a probabilistic sensitivity analysis. Assuming normally-distributed errors around the reported estimates of RSV-associated SARI hospitalizations, we fitted the distributions to the median and 95% credible intervals from our previous model [6,10]. We generated 10,000 samples per age group and used these in estimating the median and 95% credible intervals of the benefit endpoints.

#### Efficacy and Waning of Vaccine Protection

Our previous model [10] assumed that vaccine efficacy was either constant or waned exponentially over its duration of protection, based on fitting to trial data available at that time. Given the observed vaccine efficacy waning in the trial [7,13], we re-estimated vaccine efficacy and waning of protection from the trial data using the following Bayesian framework. We assumed that vaccine efficacy against severe and less severe RSV-associated medically-attended (MA) lower respiratory tract infections (LRTIs) [13,19] wanes following an Erlang-2 distribution. We sampled parameters to reflect a consistent rate of waning protection across both outcomes, while allowing the initial strength of protection to potentially differ for each outcome. To characterize waning vaccine-derived immunity after birth, we fitted the model jointly to the trial observations for severe and less severe RSV-associated MA-LRTIs [13] using Markov chain Monte Carlo (MCMC). We used the observations from 0 to 180 days after birth grouping into 30-day intervals as presented by Munjai et al [13]. The credible intervals for the number of averted deaths include both the uncertainty from estimated RSV CFR and that of the fitted vaccine efficacy. Further details are described in SA1 Appendix.

### Risks

Fatal outcomes among preterm births were rare in the trial: in the South African component of the trial, two neonatal deaths in the intervention arm and two in the placebo arm were reported during follow-up [1]. To estimate excess neonatal deaths potentially associated with vaccination, we instead used GA-specific neonatal mortality estimates from a South African longitudinal birth cohort study [2] and combined those with the probability of delivery at a specific gestational age in the trial. We constructed distributions of GA at delivery for vaccinated and unvaccinated mothers using the trial data. We then calculated expected excess deaths using differences in GA-specific delivery proportions among vaccinated and unvaccinated mothers, and neonatal mortality estimated from Zar et al [2].

#### Gestational Age-Speciffic Delivery Risk of Vaccinees

The difference in delivery proportions at each GA among vaccinated and unvaccinated mothers was calculated from the trial observations [1]. The primary analysis was conducted using all GA at birth data as observed in the South African component of the trial. We conducted scenario analyses by excluding early preterm births (< 34 weeks) in the trial. We also estimated the risk when using only trial data for live births born to mothers vaccinated at 27-36 gestational weeks in the South African component. This analysis attempts to simulate what might happen if vaccination was only delivered within this GA window (not just by excluding early preterm infants).

To account for the uncertainty in GA distributions, we bootstrapped (10,000 iterations) the number of births by trial arm in South Africa, focusing on gestational ages at which infants were born.

### Gestational Age-Speciffic Neonatal Mortality

Using a Bayesian framework, we estimated GA-specific neonatal mortality risk in South Africa from data from the Drakenstein Child Health Study (DCHS) [2], a longitudinal birth cohort study, in which from 2012 to 2015 women were enrolled in their second trimester of pregnancy from peri-urban health facilities. Recorded outcomes include live births as well as neonatal deaths stratified by gestational age, specifically <28 weeks, 28, 29, 30, 31, 32, 33, 34, 35, 36, and 37+ weeks gestation.

We fitted an Erlang-2 distribution to data on neonatal mortality at 28-36 weeks to obtain a smooth function of GA-specific neonatal mortality. Outside this range, because of lack of disaggregated data, we assumed that for births born before 28 weeks, the neonatal mortality was the same as that at 27 weeks. For infants born after 36 weeks, instead of assuming uniform distribution of the data among the subgroup, we assumed the number of births follows the distribution in the South African component of the phase 3 trial [1] and all the deaths observed in the South African cohort study were infants born at 37 weeks, i.e., the earliest GA in that subgroup. We also assumed that neonatal mortality risk was constant from 37 weeks onwards.

We fitted the function to the births and deaths using MCMC. We also conducted a scenario analysis using pooled birth and death data from the Vulnerable Newborn Measurement Collaboration (VNMC), a study combining population-based data from 15 LMICs from 2000 to 2017 [20] instead of the aforementioned assumption for constant neonatal mortality for infants born after 36 weeks gestation as neonatal mortality likely varies by gestational age (SA4 Appendix).

GA dating in DCHS was performed by second-trimester ultrasonography which is consistent with the trial where most participants GA was established by the second trimester ultrasound [2]. We bootstrapped gestational ages in Zar et al. assuming GA assessment by second-trimester ultrasonography has an accuracy of ±14 days [21]. This was assumed to be uniformly distributed. The details are described in SA5 Appendix.

### Scenario Analysis

Extremely and very preterm births (born before 32 weeks gestation) were rare (<0.5% of all births) in both placebo and intervention arms of the trial [1]. We performed scenario analyses to investigate how influential these small numbers of early preterm births were. We estimated excess neonatal mortality in South Africa under different assumptions regarding data from the trials: (i) excluding the earliest birth in each trial arm (i.e., 27 weeks in the intervention arm and 30 weeks in the placebo arm), and (ii) excluding the five earliest births in each trial arm.

We performed a scenario analysis with the highly optimistic assumption that vaccine efficacy remains 80% for the first year of life.

## Results

### Infant deaths averted through vaccination

The modelled efficacy against severe RSV-associated MA-LRTIs was 87% (95%Crl: 67, 98) on the first day of life and that against less severe RSV-associated LRTI was 65% (95%Crl: 45, 85). Efficacy waned to 10% (95%Crl: 1.6, 47) and 7.5% (95% Crl: 1.2, 32) a year following birth, respectively (Fig 1).

**Fig 1.**
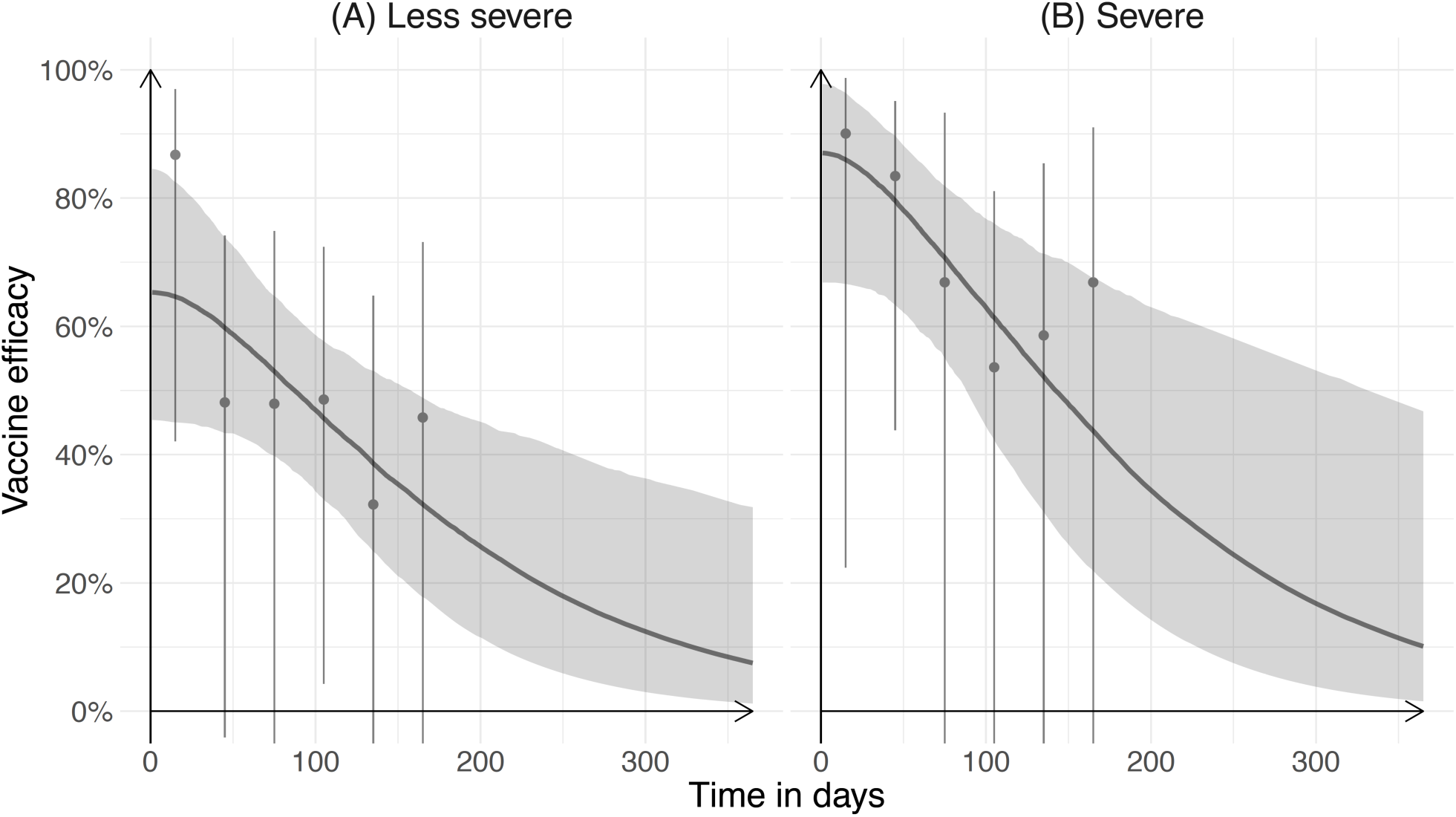
Vaccine efficacy of RSVpreF against RSV-associated (A) less severe and (B) severe MA-LRTIs during the first year of life. Vaccine efficacy against RSV-associated severe MA-LRTIs observed in the trial is shown as gray dots together with binomial 95% confidence intervals. Modelled efficacy is shown as a gray line with gray-shaded 95% credible intervals.

Using this vaccine efficacy in the impact model, we estimated that maternal RSV vaccination would prevent 31 (95%Crl: 27, 35) RSV-associated infant deaths per 100,000 live births born to vaccinated mothers in South Africa.

### Neonatal deaths potentially associated with vaccination

We modelled neonatal mortality risk as a function of GA week using data from the large South African birth cohort study [2], we estimated that in South Africa neonatal mortality within the first 28 days of life if born at 37 weeks of gestation or later was 370 (95%Crl: 170, 800) per 100,000 live births (Fig 2A). Neonatal mortality increased to 3,600 (95%Crl: 2,000, 6,100) per 100,000 live births if born at 32 weeks and 24,000 (95%Crl: 11,000, 42,000) at 27 weeks. Combining the modelled neonatal mortality and the observed GA at birth in the trial arms (Fig 2C), we estimated that the excess neonatal mortality risk associated with preterm birth risk was 44 (95%CrI: −43, 210) per 100,000 live births.

**Fig 2.**
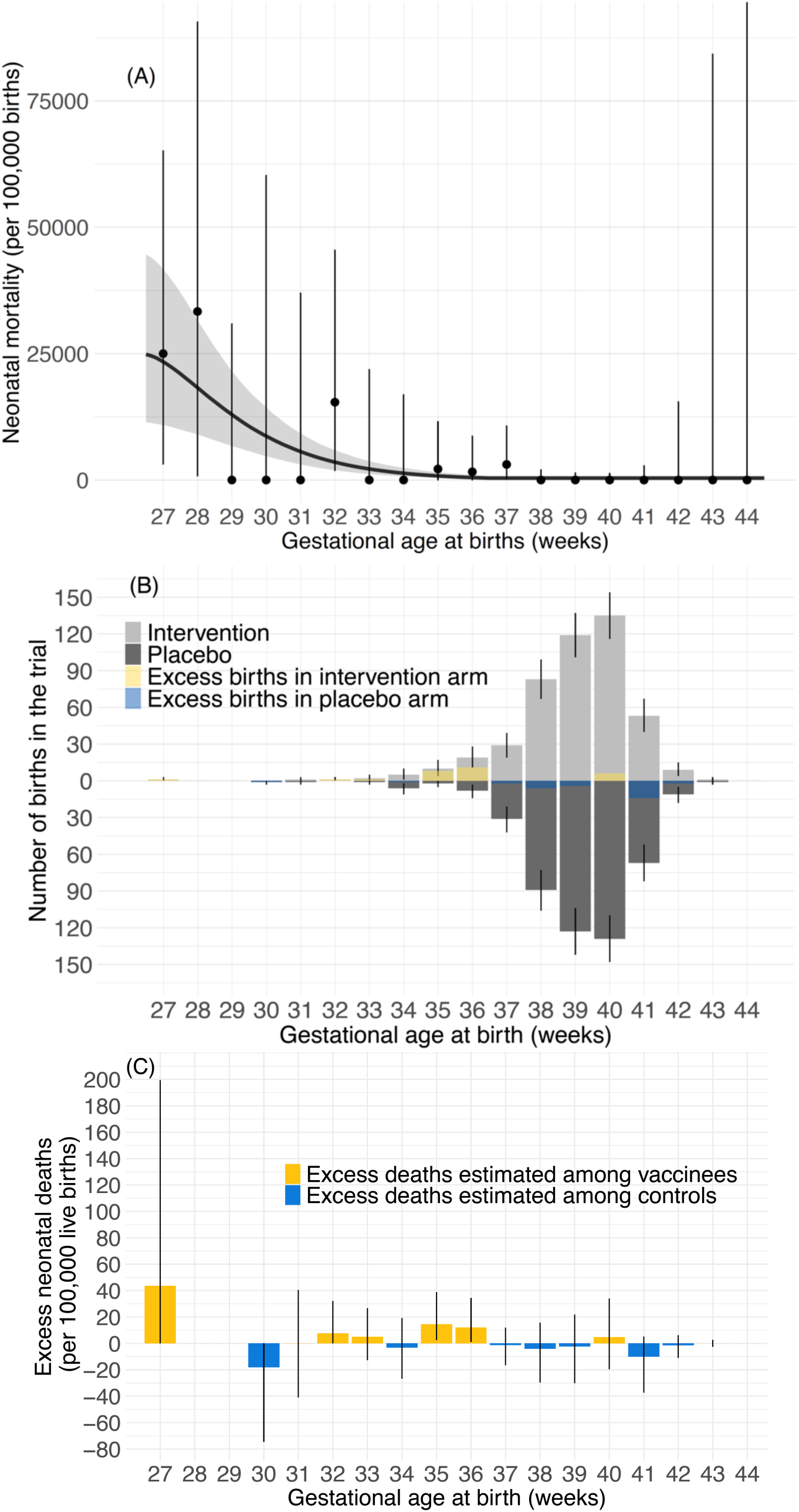
Gestational age (GA)-specific estimates of neonatal mortality in South Africa. **(A) Neonatal mortality per 100,000 live births by GA at birth from Zar et al [2].** Observed mortality is shown as dots together with binomial 95% confidence intervals. Modelled neonatal mortality is shown as a gray curve line with gray-shaded 95% credible intervals. **(B) GA-specific births born to mothers vaccinated or given placebo at 24-36 GA weeks in the South African component of the trial by trial arm**. Bootstrapping trial birth observations, light and dark gray bars indicate the median number of births in the intervention and placebo arms, respectively. The yellow and blue bars show the difference in medians of numbers of births at each GA. Yellow bars indicate a higher median in the intervention arm, while blue bars indicate a higher median in the placebo arm. Error bars show 95% credible intervals of the medians. **(C) Estimated GA-specific excess neonatal deaths per 100,000 live births born to mothers vaccinated or given placebo at 24-36 GA weeks**. Bars show estimated excess neonatal deaths at each GA. If modelled deaths are larger among newborns born to vaccinated mothers, the bars are colored yellow. If modelled deaths are larger among newborns born to unvaccinated mothers, the bars are colored blue. Error bars show 95% credible intervals.

We applied the model-estimated neonatal mortality risks in the primary analysis to the number of live births observed in the two trial arms in the South African component. We estimated that 2.0 and 2.2 neonatal deaths would have occurred among infants born to unvaccinated and vaccinated mothers respectively. This compares to 2 observed neonatal deaths in both the placebo and intervention trial arms. Similarly, applying the model-estimated RSV-associated risk for infant deaths to the number of live births in each South African trial arm, we estimated 0.24 and 0.095 infant deaths among infants born to unvaccinated and vaccinated mothers, respectively. This compares to no RSV-associated infant deaths observed in either trial arm.

However, when we restricted our analyses to include only trial data for mothers vaccinated at 27 GA weeks onwards, 111 infants in the intervention arm and 131 in the placebo arm were excluded. We then estimated the risk to be −24 (95%Crl: −120, 31) neonatal deaths per 100,000 live births born to vaccinated mothers; i.e., a net reduction in neonatal mortality based on differences in preterm birth among trial arms (Fig 3B).

**Fig 3.**
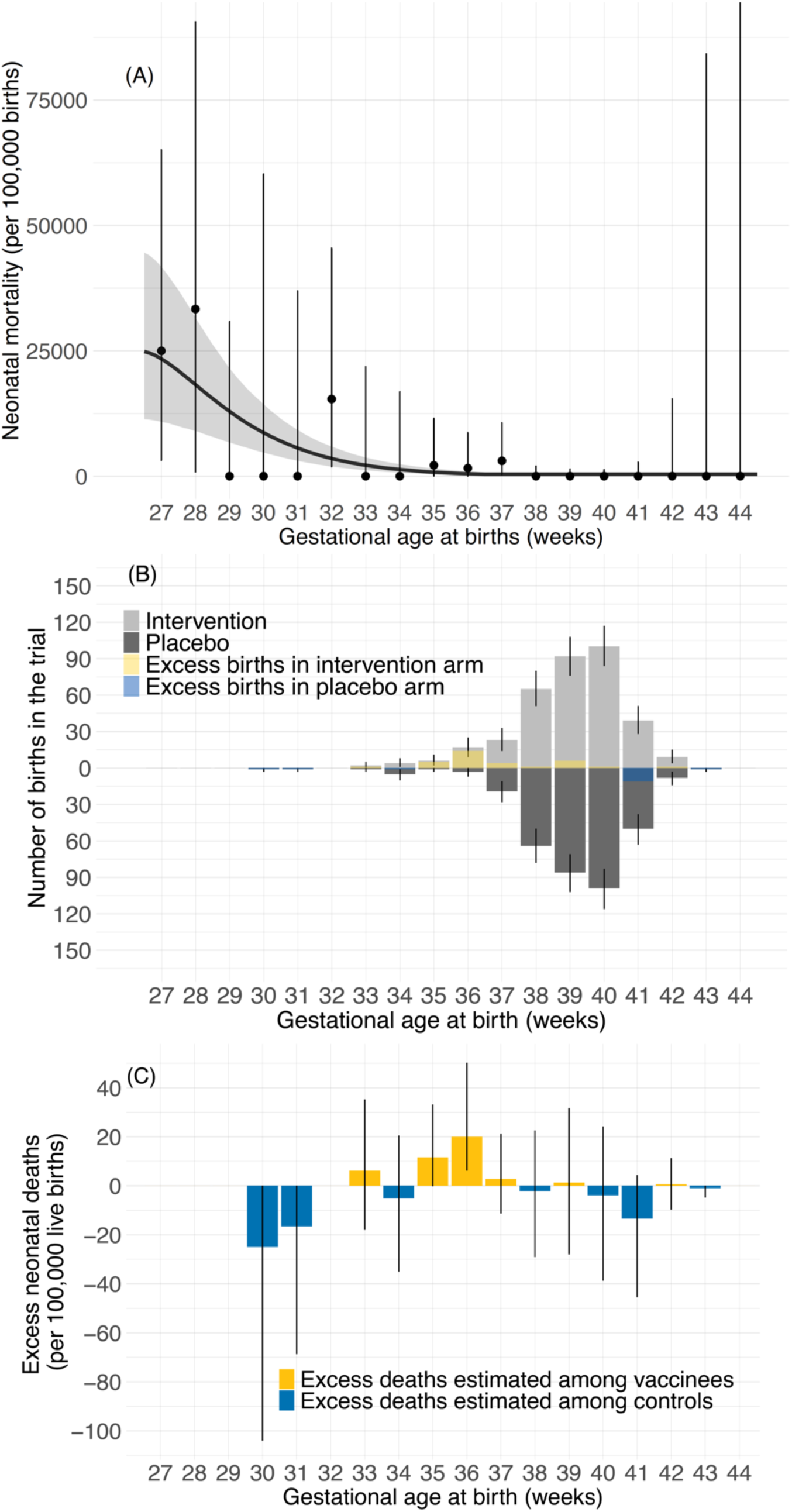
Gestational age (GA)-specific estimates of vaccination-associated neonatal mortality in South Africa using trial outcomes of live births born to mothers vaccinated or given placebo at 27-36 GA weeks in the South African component. **(A) Neonatal mortality per 100,000 live births by GA at birth from Zar et al** [2]. Observed mortality was shown as dots together with 95% confidence intervals. Modelled neonatal mortality is shown as a gray curve line with gray-shaded 95% credible intervals. (**B) GA-specific births born to mothers vaccinated or given placebo at 27-36 GA weeks in the South African component of the trial by trial arm**. Bootstrapping trial birth observations, light and dark gray bars indicate the median number of births in the intervention and placebo arms, respectively. The yellow and blue bars show the difference in medians of numbers of births at each GA. Yellow bars indicate a higher median in the intervention arm, while blue bars indicate a higher median in the placebo arm. Error bars show 95% credible intervals of the medians. (**C) Estimated GA-specific excess neonatal deaths per 100,000 live births born to mothers vaccinated or given placebo at 27-36 GA weeks**. Bars show estimated excess neonatal deaths at each GA. If modelled deaths are larger among newborns born to vaccinated mothers, the bars are colored yellow. If modelled deaths are larger among newborns born to unvaccinated mothers, the bars are colored blue. Error bars show 95% credible intervals.

### Comparison of benefit and risk

For South Africa, we estimated that with vaccination in 24-36 GA weeks, there may be an excess of 13 (95%Crl: −74, 180) neonatal deaths per 100,000 live births associated with maternal RSV vaccination (Table 1). For every infant (between birth and 12 months) saved through protection against RSV by maternal vaccination, there may be 1.4 (95%CrI: −1.4, 7.0) excess neonatal deaths associated with potentially vaccine-associated preterm birth. In 41% of our simulations, the estimated benefit exceeded the estimated risk. In 22% of our simulations, at least five infant deaths were prevented for each neonatal death associated with vaccine-associated preterm birth (Table 1, Fig S1).

**Table 1.** Risks and benefits by scenario. The risk is measured by excess neonatal deaths due to vaccine-associated preterm birth per 100,000 live births born to vaccinated mothers, and the benefit is measured by the vaccine-preventable RSV-associated infant deaths per 100,000 live births born to vaccinated mothers. ‘Base case’ presents the estimates using all births in the South African component of the trial. ‘27-36 GA weeks vaccination’ presents the estimate using trial outcomes of infants born to mothers vaccinated (or given placebo) at 27-36 weeks. ‘Earliest birth removed’ and ‘Earliest 5 births removed’ present the estimates using births in the South African component without the first earliest birth in each arm (i.e., 27 weeks in the intervention arm and 30 weeks in the placebo arm), and estimates using births in South African component without the 5 earliest births in each arm, respectively. For the four scenarios, the benefits are estimated using all births in South Africa in the trial (i.e., vaccinated at 24-36 weeks).

| **Scenario** | **Risk**<br>(excess neonatal deaths due to vaccine-associated preterm birth per 100,000 live births born to vaccinated mothers)<br>Median (95%CrI) | **Benefit**<br>(vaccine-preventable RSV-associated infant deaths per 100,000 live births born to vaccinated mothers)<br>Median (95%CrI) | **Net mortality**<br>(= risk – benefit) per 100,000 live births born to vaccinated mothers<br>Median (95%CrI) | **Risk-benefit ratio** (Excess neonatal deaths (risk) per 1 infant saved by vaccination (benefit))<br>Median (95%CrI) | Percentages of simulations where the benefit exceeds the risk | Percentages of simulations where the benefit exceeds 5 times the risk |
| --- | --- | --- | --- | --- | --- | --- |
| Base case | 44 (-43, 210) | 31 (27, 35) | 13 (-74, 180) | 1.4 (-1.4, 7.0) | 41% | 22% |
| 27-36 GA weeks vaccination | -24 (-120, 31) | 31 (27, 35) | -55 (-150, 0.17) | -0.76 (-3.9, 1.0) | 98% | 84% |
| Scenario analysis |  |  |  |  |  |  |
| Earliest birth removed | 21 (-20, 78) | 31 (27, 35) | -10 (-51, 47) | 0.69 (-0.67, 2.5) | 67% | 23% |
| Earliest 5 births removed | 15 (-0.42, 39) | 31 (27, 35) | -16 (-32, 8.4) | 0.48 (-0.014, 1.3) | 92% | 15% |

With risk estimated using data for infants born to mothers vaccinated or given placebo at 27-36 GA weeks, the estimated benefit exceeds the risk by 55 (95%Crl: −0.17, 150). In 98% of simulations, the estimated benefit exceeds the estimated risk, and in 84% of simulations, the benefit exceeded the risk by more than a factor of five (Table 1, Fig 4).

**Fig 4.**
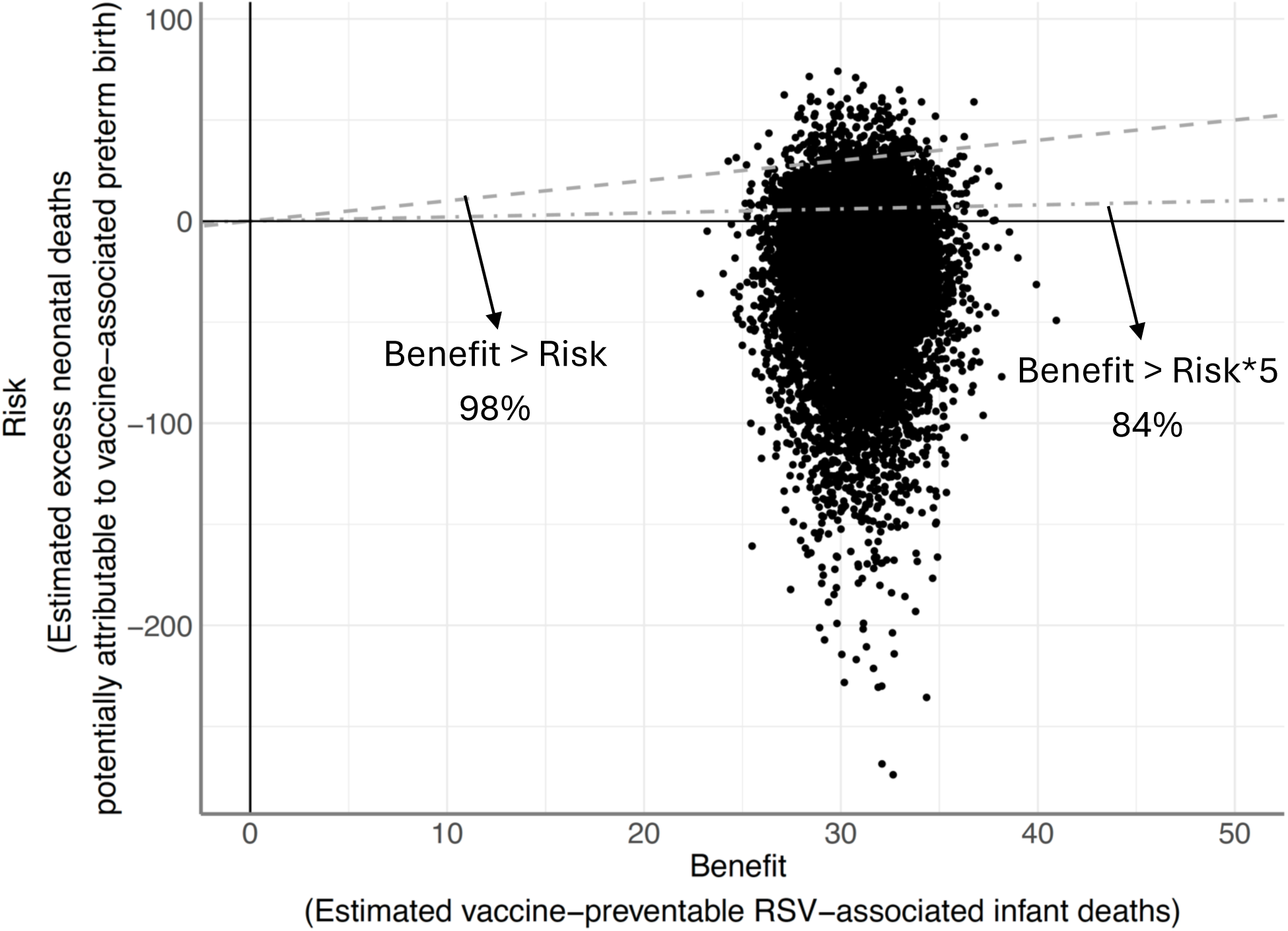
Estimated risks of RSVpreF in South Africa using trial birth outcomes born to mothers vaccinated (or given placebo) at 27-36 GA weeks compared to estimated benefits. The risk is measured by the excess neonatal deaths potentially attributable to vaccine-associated preterm birth estimated using trial birth data of infants born to mothers vaccinated (or given placebo) at 27-36 GA weeks (subset of data). The benefit is measured by the number of vaccine-preventable RSV-associated infant deaths in less than 1-year-old infants using vaccine efficacy estimated from trial data of infants born to mothers vaccinated at 24-36 GA weeks (full dataset). Dots indicate posterior samples of the estimated risk and benefit. Dots below the dashed line indicate that benefit exceeds risk in that simulation (i.e., more than one life would be saved per every one life lost). Dots below the dot-dash line indicate that the benefit exceeds five times the risk in that simulation (i.e., more than five lives would be saved per every one life lost). Percentages besides the lines indicate the percentage of simulations that exceeds the benefit-risk ratio of 1:1 and 5:1, respectively.

### Scenario Analysis

#### Exclusion of early births

In our modelled results, a small number of very early preterm births in the trial data substantially influenced the estimated benefit-risk ratio (Fig 2B and 2C). When the earliest birth in each arm (i.e., a 27-week birth in the intervention arm and a 30-week birth in the placebo arm) was excluded, the excess neonatal deaths attributable to preterm birth was 21 (95%Crl: −20, 78) per 100,000 live births born to vaccinated mothers, and 0.69 (95%CrI: −0.67, 2.5) excess neonatal deaths may be associated with vaccination per every one infant life saved through protection against RSV through vaccination (Table 1). Lives saved exceed associated deaths in 67% of the simulations.

Excluding the five earliest births in each arm, the excess neonatal deaths would be 15 (95%CrI: −0.42 39) per 100,000 live births born to vaccinated mothers and 0.48 (95%CrI: −0.014, 1.3) excess neonatal deaths may be associated with vaccination per every one infant life saved through vaccination. Lives saved exceed associated deaths in 92% of our simulations.

#### Vaccine efficacy

If vaccine efficacy is assumed to remain constant at 80% throughout the first year of life, the estimated benefit in South Africa increases, but the point estimate would not substantially outweigh that of the risk (SA3 Appendix). Further details of the sensitivity analyses are described in the Supplement.

## Discussion

If vaccination is introduced at 24-36 GA weeks, the estimated benefit of maternal vaccination through reduction in RSV-associated infant mortality is unlikely to substantially outweigh the potential risk of increased neonatal mortality due to vaccine-associated preterm birth in South Africa. However, there is considerable uncertainty around our modelled estimate of vaccine-related neonatal mortality risk. We also estimate that if vaccination is introduced at 27-36 GA weeks, the mortality benefit is likely to outweigh the risk. These findings are based on the assumption that the observed increased preterm birth was related to RSVpreF as observed in the South African component of the trial, and that this increased preterm birth risk leads to neonatal deaths based on GA-specific risks derived from a South African birth cohort study. The findings have been presented at the SAGE meeting in September 2024, and SAGE ultimately recommended RSV vaccination in the third trimester of pregnancy, as defined by the local context which in most countries is 28 GA weeks onwards [22]. Again, the observed association may not be causal, i.e., vaccination may not have caused the preterm births.

Our conclusions are largely dependent on a relatively small number of early preterm infants in the trial, and we find that the point estimate of the benefit-risk ratio could reverse if we exclude these early preterm infants in the trial. It is hard to compare these benefit-risk ratio with other vaccines’ ratios, which are context-specific [23–25]. In the scenario analysis, we ran one scenario in which the risk and benefit of vaccination were weighted equally. Meanwhile, we also ran another scenario in which the risk is weighted five times the benefit, given that debates regarding decision-making in vaccination; i.e., people may put more emphasis on the risk of vaccination than on the benefit [23,26,27]. The results are also subject to uncertainty surrounding limited data on mortality of early preterm infants. Furthermore, by applying the model-estimated neonatal mortality risks to the number of live births in the South African component of the trial, the findings indicate that expected excess deaths either in the benefit or the risk were too small to be detected among the small number of live births in the South African component of the trial, in which there was no increase in deaths based on vaccination. As SAGE noted, the vaccine trials had too few participants from LMICs to estimate the full impact of the intervention in these settings [22,28]. Given this under-representation, the observed numerical imbalance may simply be a Type I error, or could reflect a (currently unknown) biological mechanism. While our study cannot determine which is more likely, it extrapolates South African trial birth outcomes assuming the effect is genuine.

Maternal RSV vaccination with restricted GA windows has been licensed by several regulatory authorities in high-income countries (HICs) [17,29]. Notably, licensure indications for RSV immunisation vary among HICs; e.g., EMA allows for maternal vaccination from 24 GA weeks onwards. In South Africa, the maternal vaccine has been licensed for use in pregnant women between 28 and 36 GA weeks, and the NITAG has recommended the vaccination during this period, i.e., from the third trimester onwards. Although there remain challenges to practicing this in LMICs (e.g., there is considerable uncertainty around GA assessment [30], timing of attendance to antenatal care [31], etc.), our analyses indicate that with vaccination from 27 weeks onward, the benefits may outweigh the risks. Our analyses and the subsequent SAGE recommendation support decision-makers in LMICs in introducing maternal RSV vaccination in their countries. Also, post-licensure surveillance is needed to monitor the association carefully in order to address concerns about potential causality between preterm birth and vaccination. There is ongoing post-marketing surveillance to assess potential adverse outcomes including preterm birth among vaccinees in early-introducing countries (e.g., U.S., Argentina, U.K.) [32,33]. Moreover, a multisite phase 4 study is planned in Africa that will evaluate preterm births.

Our study design had several limitations. Firstly, our analysis focuses solely on mortality due to the impact of RSV and preterm birth. However, the burden of both conditions can extend beyond mortality. For instance, our previous analysis estimated that maternal vaccination would reduce RSV-associated hospitalizations in South Africa by 24.2% (95%CrI: 18.7, 28.6) and RSV-associated deaths by 27.4% (95%CrI: 21.6, 32.3) [10]. Both RSV-associated LRTI during early childhood and preterm birth have also been linked to long-term consequences [34–36]. Although most of the total disease burden, as measured through disability-adjusted life years, is due to deaths, evaluating non-fatal and long-term outcomes would provide further refinement to the estimated benefit-risk ratio from vaccination. We also used estimates of reductions in RSV-associated infant deaths based on RSV disease burden in 2011-16 in South Africa [6]. Also, we did not include analysis of the potential secondary benefits of prevention of severe RSV disease in infants through freeing up resources for other conditions (e.g., more availability of hospital beds, etc), enabling reduction of mortality from other treatable causes [37]. In addition, we estimated benefits using efficacy estimated from all births in the South African component of the trial (i.e., births born to mothers given intervention or placebo at 24-36 GA weeks. Meanwhile, for the 27-36 GA weeks analysis, we estimated risks using data from the subset of trial births: i.e., we used 27-36 GA weeks vaccinated (or given placebo) dataset for the 27-36 weeks analysis, while using 24-36 weeks dataset for the 24-36 weeks analysis because of limited data availability. This assumes that vaccine efficacy is consistent between mothers vaccinated at 24-36 weeks and those vaccinated at 27-36 weeks.

Another limitation is that we used the baseline preterm birth risk from the placebo arm of the trial, which is substantially lower than the overall preterm birth risk in South Africa [38,39]. For instance, Ohuma et al. estimated that preterm birth rate in 2020 is 13 (95%CI: 9.2, 18) per 100 live births in South Africa [39].

In addition, another limitation is that our conclusions were very sensitive to the outcomes associated with early preterm infants, but as the South African cohort data aggregated the neonatal mortality risk before 28 weeks, we did not know the exact GA-specific neonatal mortality risk before 28 weeks and instead assumed a constant risk. Moreover, our estimates of GA-specific neonatal mortality are based on a study conducted between 2012-15 [2], and using more recent neonatal survival rates may change our conclusions.

It is currently not established nor understood whether the observed association between vaccination and preterm birth is genuine or causal [12,40].The numerical imbalance in preterm birth in the trial was only statistically significant in South Africa and occurred predominantly at peaks of the delta and omicron waves of SARS-CoV-2 [13]. A similar preterm birth imbalance was observed in a trial of another pre-F maternal vaccine also undertaken during the Covid-19 pandemic [14]; however, no imbalance in preterm births was observed in a pre-pandemic trial of another maternal RSV vaccine that enrolled over half of the participants in South Africa [41]. Moreover, in the MATISSE trial most infants were born more than 30 days after vaccination, and there was no temporal relationship or proposed biological mechanism between the vaccination and preterm birth [17].

Post-licensure surveillance is needed to clarify if RSVpreF and preterm birth are associated [13,14,42], however, it was out of the scope of our analysis. Our analysis did not consider some other key outcomes that are potentially important, including severe RSV disease associated with preterm birth [43], stillbirths, and other fetal deaths [12], or seasonal and other temporal variations in RSV incidence [6] and preterm birth risk [17].

Our study illustrates the potential importance of the observed imbalance in preterm birth following maternal RSV vaccination at broader GA vaccination windows. However, we also show that any potential risk could be largely mitigated by changing vaccine eligibility to begin in the third trimester. The first long-awaited maternal RSV vaccine has recently been recommended by WHO SAGE for use in the third trimester of pregnancy and will likely being globally available in the next few years. Post-marketing surveillance is important to obtain further evidence about its safety and effectiveness when used in real-world settings.

## Data Availability

This analysis uses data presented at the SAGE meeting in September 2024, which includes data shared by Pfizer from the Phase 3 MATISSE trial. Pfizer did not participate in the analysis of such data nor did Pfizer have any role in the conclusions drawn from the analysis presented by SAGE. Data from the Drakenstein Child Health Study were obtained from Heather Zar (personal communication); some of these data (on the total number of births) were available in the publication. Some additional data (on the number of births by gestational age and the number of neonatal deaths) was not published but was shared in confidence at the SAGE meeting; this can be obtained through personal communication with the study team as we do not own the data. Data analysis codes used in this study are available at https://github.com/ayakamon/BR-RSV-MV.

## Availability of data

This analysis uses data presented at the SAGE meeting in September 2024 [1], which includes data shared by Pfizer from the Phase 3 MATISSE trial. Pfizer did not participate in the analysis of such data nor did Pfizer have any role in the conclusions drawn from the analysis presented by SAGE. Data from the Drakenstein Child Health Study [2] were obtained from Heather Zar (personal communication); some of these data (on the total number of births) were available in the publication [2]. Some additional data (on the number of births by gestational age and the number of neonatal deaths) was not published but was shared in confidence at the SAGE meeting; this can be obtained through personal communication with the study team as we do not own the data. Data analysis codes used in this study are available at https://github.com/ayakamon/BR-RSV-MV.

## Ethical approvals

This study has received ethical approval from the London School of Hygiene & Tropical Medicine Ethics Committee (Ref: 29955).

## Funding

AM was funded by the Japanese Ministry of Education, Culture, Sports, Science and Technology through the Doctoral Program for World-leading Innovative & Smart Education as part of the NU-LSHTM Joint PhD Programme for Global Health (https://www.mext.go.jp/en/policy/education/highered/title02/detail02/1373919.html). AE is supported by the Japan Science and Technology Agency (JPMJPR22R3, https://www.jst.go.jp/EN/), Japan Society for the Promotion of Science (JP22K17329, https://www.jsps.go.jp/english/), and Japan Agency for Medical Research and Development (JP223fa627004, https://www.amed.go.jp/en/). SF is funded by the Einstein Foundation Berlin as an Einstein BUA Strategic Professor (EPP-BUA-2022-697, https://www.einsteinfoundation.de/en/). The funders had no role in study design, data collection and analysis, decision to publish, or preparation of the manuscript. HJZ was funded by the Bill & Melinda Gates Foundation (grants OPP1017641 and OPP1017579, https://www.gatesfoundation.org/) for the Drakenstein Child Health study.

AM’s travel for collaborative visits for this project was supported by funding from Bill & Melinda Gates Foundation (INV-069494, https://www.gatesfoundation.org/). MJ, SIL, and SRP were also supported by funding from Bill & Melinda Gates Foundation (INV-069494, https://www.gatesfoundation.org/). This funder had roles in the study design, and data collection through its membership on the maternal RSV vaccine benefit-risk advisory group, but no role in data analysis, decision to publish, or preparation of the manuscript.

## Competing interests

SAM institution has received funding from Pfizer, GSK, MSD and AstraZeneca for research on RSV vaccines and RSV monoclonal antibodies. PB institution has received public-private partnership funding through the European Union IMI/IHI Respiratory Syncytial Virus Consortium in Europe (RESCEU) and Preparing for RSV immunisation and surveillance in Europe (PROMISE) projects (ceased in 2023). HJZ has received funding from Pfizer, MSD, and Sanofi for studies of RSV vaccines and monoclonal antibodies and serves on the Data Safety Management Board for Moderna RSV maternal vaccine and advisory boards to MSD and Pfizer.

## Abbreviations

CFR: case fatality rate
CI: confidence interval
Crl: credible interval
DCHS: the Drakenstein Child Health Study
EMA: European Medicines Agency
FDA: the US Food and Drug Administration
GA: gestational age
LMIC: low- and middle-income countries
LRTI: lower respiratory tract infection
MCMC: Markov chain Monte Carlo
MA: medically-attended
RSV: Respiratory syncytial virus
RSVpreF: RSV prefusion F protein vaccine
SAGE: Strategic Advisory Group of Experts on Immunization
SARI: severe acute respiratory illness
VNMC: the Vulnerable Newborn Measurement Collaboration
WHO: the World Health Organization

## Author Contributions

Conceptualization: MJ, AM

Data curation: AM

Formal analysis: AM

Investigation: All authors

Funding acquisition: MJ

Methodology: AM, MJ, SF, AE, SRP, SIL

Software: AM, SF, AE, SIL

Validation: All authors

Supervision: MJ, SF, AE

Writing – original draft preparation: AM

Writing – review and editing: All authors

All authors provided final approval of the version to be published and agreement to be accountable for all aspects of the work in ensuring that questions related to the accuracy or integrity of any part of the work are appropriately investigated and resolved.

## Acknowledgments

We are grateful to James Nokes, Padmini Srikantiah and Angela Guo for helpful input throughout the project. We are also grateful to Clare Cutland for information on the GA dating in LMICs. The findings and conclusions contained within are those of the authors and do not necessarily reflect positions or policies of the World Health Organization.

## Supporting information

### A1 Estimates of waning efficacy

#### Model outline

Observed disease outcomes in trial suggested vaccine efficacy wanes after birth [7,13]. We modelled VE_est(t) vaccine efficacy at time t after birth as the product of V0 initial value of efficacy and *w(t)* the function of protection decay after birth assuming it follows an Erlang-k distribution in which both the shape and scale parameters can be estimated [19].

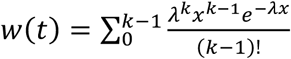

We estimated the protection waning by fitting the modelled outcomes to the observed outcomes in the trial. The incidence in the intervention arm at time t after birth *I_v(t)* was estimated using modelled efficacy and the incidence in the placebo arm *I_p(t)*.

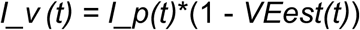

Since the trial outcomes were grouped into 30-day interval from birth to 180 days after birth, the means of the estimated vaccine efficacy during 30-day period were used to calculate the number of infected individuals in intervention arm *I_v (t)* during that period.

*C* modelled outcomes (i.e., individuals newly counted as that disease outcome during that period) in the intervention arm were fitted to the RSV disease outcomes in the trial.

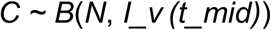

In the intervention arm*, N* is the total number of individuals in the arm, and *I_v (t_mid)* is the incidence in the intervention arm in the middle of that period assuming incidence is constant during that period. We assume that vaccine efficacy against RSV disease outcomes with different severities wanes at the same rate (λ in Equation (1)) but the initial values *V0* are severity-dependent.

The Bayesian model was implemented using a MCMC in R. Data analysis codes are published on https://github.com/ayakamon/BR-RSV-MV.

#### Overview of data

We used the final outcome of the trial (Table S1), in which the vaccine showed efficacy against RSV disease in infants born to vaccinated mothers [13]. We calculated the incremental efficacy from the data regarding cumulative counts of severe RSV-Positive MA-LRTIs (Table S1A) and less severe RSV-Positive MA-LRTIs (Table S1B) from 0 to 180 days after birth in each trial arm across the entire study.

**Table S1.** Cumulative number of trial outcomes by 30-day interval from 0 to 180 days after birth regarding (A) severe RSV-Positive MA-LRTIs and (B) less severe RSV-Positive MA-LRTIs [13].

| Time after birth (days) | RSVpreF 120µg | Placebo |
| --- | --- | --- |

|  | N = 3585 | N = 3563 |
| --- | --- | --- |
| 0 - 30 | 1 | 10 |
| 30 - 60 | 4 | 28 |
| 60 - 90 | 6 | 34 |
| 90 - 120 | 13 | 49 |
| 120 - 150 | 18 | 61 |
| 150 - 180 | 21 | 70 |

| Time after birth (days) | RSVpreF 120µg<br>N = 3585 | Placebo<br>N = 3563 |
| --- | --- | --- |
| 0 - 30 | 2 | 15 |
| 30 - 60 | 14 | 38 |
| 60 - 90 | 25 | 59 |
| 90 - 120 | 40 | 88 |
| 120 - 150 | 55 | 110 |
| 150 - 180 | 67 | 132 |

#### Fitted results

For vaccine waning, we compared the exponential model in the previous work [10] to an Erlang-2 distribution, and chose Erlang-2 distribution as it fit better to the trial outcomes. To estimate RSV-associated infant deaths averted by vaccination (benefit), we used modelled efficacy against severe outcomes during the first year of life.

### A2 Scenario analyses on trial births

**Fig S1.**
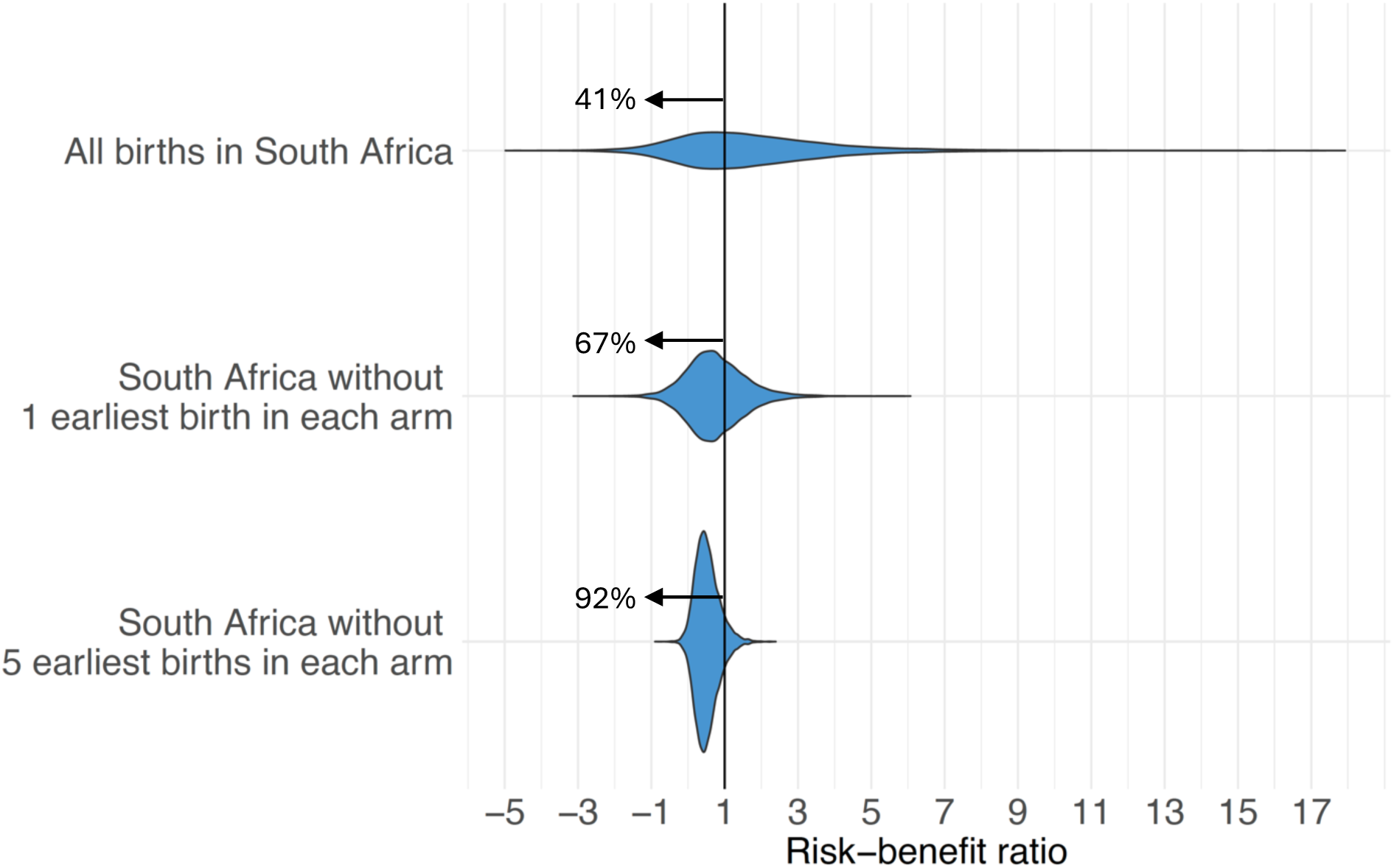
Estimated risk-benefit ratio of RSVpreF in South Africa under different scenarios on trial birth outcomes. The plots illustrate excess neonatal deaths potentially associated with vaccination per every one infant under 1 year saved through vaccination (i.e., the ratio between the risk and benefit). The risk is measured by the excess neonatal deaths potentially attributable to vaccine-associated preterm birth per 100,000 live births born to vaccinated mothers and uses bootstrapped outcomes with 10,000 iterations. The benefit is measured by the vaccine-preventable RSV-associated deaths in less than 1 year-old infants per 100,000 live births born to vaccinated mothers and uses bootstrapped outcomes with 10,000 iterations. When the ratio is below 1 (black vertical line), the estimated benefits exceeds the estimated risks. The risks are estimated using data in the South African component of the trial with all births, births in the South African component of the trial without the first earliest birth in each arm (i.e., 27 weeks in the intervention arm and 30 weeks in the placebo arm), or births in the South African component of the trial without the 5 earliest births in each arm. The percentages beside each plot show the percentage of simulations when the benefit exceeds the risk.

**Fig S2.**
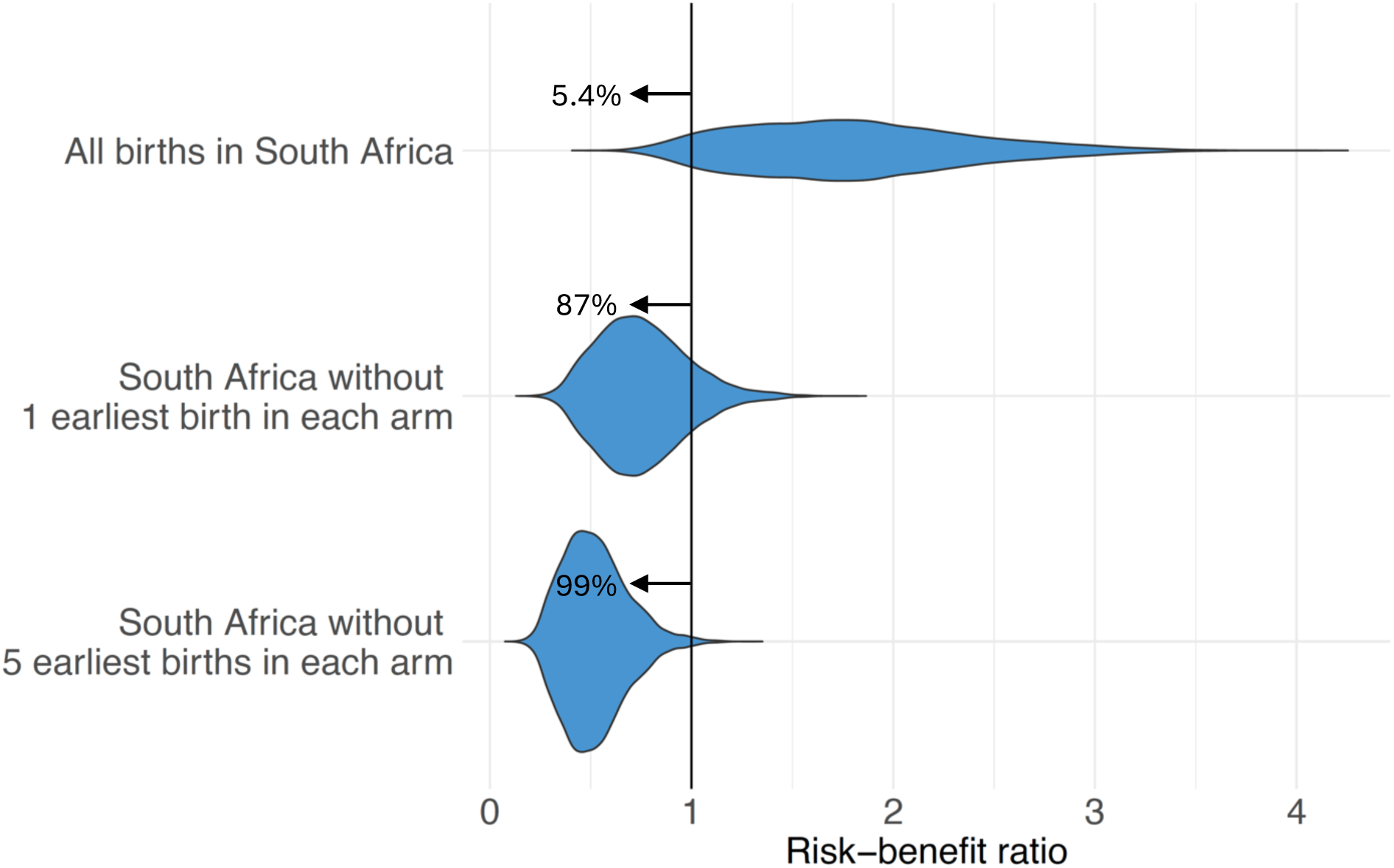
Estimated risk-benefit ratio of RSVpreF in South Africa under different scenarios on trial birth outcomes without incorporating uncertainty in trial outcomes. The risk is measured by the excess neonatal deaths potentially attributable to vaccine-associated preterm birth and uses the trial birth outcomes without bootstrapping. The benefit is measured by the vaccine-preventable RSV-associated deaths in less than 1 year-old infants per 100,000 live births born to vaccinated mothers and uses bootstrapped outcomes with 10,000 iterations. When the ratio is below 1 (black vertical line), the estimated benefits exceed the estimated benefits.The risks are estimated using all birth outcomes from the South Africa component in the trial, births in the South Africa component without the first earliest birth in each arm (i.e., 27 weeks in the intervention arm and 30 weeks in the placebo arm), and births in the South African component of the trial without the 5 earliest births in each arm.

### A3 Vaccine Efficacy

With an optimistic assumption around vaccine efficacy that vaccine efficacy against severe RSV disease remains at 80% during the first year of life without any waning, the estimated benefit in South Africa increases from the waning vaccine assumption of 31 (95%CrI: 27, 35) to 41 (95%CrI: 38, 44) vaccine-preventable RSV-associated infant deaths per 100,000 live births born to vaccinated mothers.

### A4 Neonatal mortality of neonates born after 36 weeks

Instead of assuming constant neonatal mortality of neonates born after 36 GA weeks, we conducted a scenario analysis using birth data from VNMC, a study combining population-based data from 15 LMICs [20]. We used GA-specific neonatal mortality data from VNMC for live birth born after 36 GA weeks devided by the overall neonatal mortality after 36 GA weeks from VNMC. Then we calucuated the total number of deaths per total number of live births born after 36 GA weeks from DCHS. Then we multiplied the mean mortality of live birth born after 36 GA weeks from DCHS by the standardized GA-specific neonatal mortality for each GA after 36 GA weeks from VNMC. We then multiplied the VNMC adjusted DCHS GA-specific deaths devided by live births by the bootsrapped birth outcomes in the trial to estimate the excess neonatal deaths potentially associated with vaccination. We then estimated that the excess neonatal deaths potentially associated with vaccination would be 56 per 100,000 live births born to vaccinated mothers. Compared to the risk estimates assuming that all the deaths observed in the South African cohort study were only among infants born at 37 weeks and that neonatal mortality was constant after 36 weeks, the estimated neonatal mortality is generally smaller (e.g., for infants born at 37 GA weeks, 126 per 100,000 live births). Allowing neonatal mortality risk to vary after 36 weeks (as described above) results in a slight increase in excess deaths among vaccinees compared to controls (i.e., excess deaths increased from 44 per 100,000 live births born to vaccinated mothers from the main analysis, to 56 per 100,000 live births born to vaccinated mothers).

### A5 GA dating

GA dating by ultrasonography was performed at the second trimester in the South African cohort study [2]. However, this technique is believed to be only accurate within a margin of ±14 days [21]. We can incorporate this margin of error to further increase the uncertainty around GA at birth, but this has little effect on the point estimates of the risks of vaccination, or the conclusion that benefits are unlikely to outweigh the risks of neonatal deaths if vaccination is provided between 24-36 GA weeks.

**Table S2.** Estimated excess neonatal deaths using outcomes in Zar et al. with bootstrapping on GA dating. 5 sets of results from the bootstrap are shown.

| Excess deaths per 100,000 live births born to vaccinated mothers (95%CrI) | Excess deaths per life saved through vaccination (95%CrI) |
| --- | --- |
| 49 (-33, 190) | 1.3 (-1.1, 6.2) |
| 46 (-30, 180) | 1.2 (-0.98, 5.7) |
| 49 (-37, 190) | 1.3 (-1.2, 6.2) |
| 47 (-33, 180) | 1.2 (-1.1, 5.8) |
| 53 (-35, 200) | 1.3 (-1.1, 6.7) |

